# Spatial variation in incidence of meningococcal meningitis: evidence from a large historical epidemic in Glasgow

**DOI:** 10.64898/2026.05.28.26354324

**Authors:** Gillian Stewart, Max Schroeder, Rebecca Mancy, Konstantinos Angelopoulos

**Affiliations:** School of Health and Wellbeing, University of Glasgow, Glasgow, United Kingdom; Durham University Business School, Durham University, Durham, United Kingdom; Wales Applied Virology Unit (WAVU), School of Medicine, Cardiff University, Cardiff, United Kingdom; School of Biodiversity, One Health and Veterinary Medicine, University of Glasgow, Glasgow, United Kingdom; School of Education, University of Glasgow, Glasgow, United Kingdom; Adam Smith Business School, University of Glasgow, Glasgow, United Kingdom; CESifo, Munich, Germany

**Keywords:** meningococcal meningitis, 1907 Glasgow epidemic, meningitis transmission, spatial variation in incidence

## Abstract

Large epidemics of invasive meningococcal disease are rare in temperate regions. Here, we analyse administrative data on the largely forgotten epidemic of bacterial meningococcal meningitis that occurred in Glasgow in 1907, probably the largest on record in the UK. The epidemic, predominantly confined to the city, killed around 1,000 people, had a case fatality rate of nearly 70%, and hit infants and young children the hardest. We show the rapid rise and fall in cases and the spatial distribution of incidence and mortality rates within the city. We find that within-household overcrowding was a key driver of incidence whereas between-household geographic proximity was not. We also find that the spatial distribution of disease risk during the epidemic persisted in the post-epidemic period and during a later outbreak. The findings suggest that interventions should prioritise populations in areas that have experienced higher incidence rates to mitigate the risk of future outbreaks.

## 1 Introduction

Most meningitis cases occur as sporadic and apparently independent events (Chang et al., 2012), with large epidemics of meningitis rare in temperate regions and more generally outside of the so-called African Meningitis Belt. Prior to the outbreak in Kent in 2026, recent outbreaks in the UK have been much less explosive and typically relatively small (Mahase, 2026). Historically, however, the UK has been affected by sizeable epidemics of invasive meningococcal disease, the largest of which occurred in 1906-1908, peaking in 1907, and predominantly affected Glasgow. The epidemic of meningococcal meningitis, referred to at the time as *cerebrospinal fever*, and microbiologically confirmed to be of bacterial origin, killed around 1,000 people in the city and had a case-fatality rate of nearly 70%. The epidemic had the biggest impact in Glasgow, although Belfast was also hit and medical reports and popular press coverage suggest a smaller number of cases in Liverpool and elsewhere in Ireland.

Using Medical O”cer of Health records for Glasgow and other archival sources, we analyse disease incidence and mortality rates (per 10,000 population) from the 1907 epidemic of meningococcal meningitis. We examine the time evolution of cases and deaths and their spatial pattern across the municipal areas of the city, as well as their distribution over age groups, reviewing knowledge at the time and public health responses. We then exploit the variation in incidence and mortality between municipal areas to assess the role of within-household close contact and between-household geographic proximity, controlling for spatial and socioeconomic factors. Using statistical analysis, we find that within-household close contact, as captured by measures of overcrowding and house size, was a significant predictor of incidence and mortality rates. In contrast, between-household geographic proximity, as captured by measures of housing density, did not significantly relate to incidence and mortality rates. In addition, our analysis shows an effect of socioeconomic conditions on incidence and mortality rates. Finally, we examine the persistence of the patterns of disease incidence during the fifteen years after the main epidemic wave, finding that populations in areas that showed the highest incidence rates in 1907 remained at higher risk for the following two years, and were also more strongly affected in a subsequent outbreak in 1915, over and above other observable factors. By highlighting the importance of persistence of the spatial distribution of disease risk following the epidemic, our findings contribute to ongoing discussions in 2026 about public health responses to the meningitis outbreak in Kent, UK and, more generally, to future management of meningococcal meningitis disease.

## 2 The epidemiology of the 1906-1908 epidemic in Glasgow

Prior to the epidemic of meningococcal meningitis that we analyse here, outbreaks had been seen in Europe, America, Africa and Asia, but the disease was largely unknown in the British Isles in the 1800s. The last outbreak from a previously unknown disease had been that of the so-called ‘Russian flu’ in 1891-1892, although this had impacted Glasgow relatively mildly (Smith, 1995; Schroeder et al., 2023). More generally, the context was one of sustained improvements in health in the city of Glasgow, and more generally in the UK. In the early 1900s, Glasgow was a large city that was booming economically and benefitting from sustained reductions in the mortality rate. Indeed, mortality rates in the city had fallen from 29-30 per thousand population in the late 1850s (Chalmers, 1902a) to 20 per thousand in 1905 (Chalmers, 1905). Although deaths from childhood diseases such as measles and whooping cough were still common, there had been substantial declines in mortality from the main scourges of the 19th century, including cholera, typhus, smallpox and scarlet fever.

At the time of the epidemic, there was growing recognition of the importance of reporting cases of meningococcal meningitis, but surveillance was only partially implemented. The Infectious Disease (Notification) Act 1889 (Parliament of the United Kingdom, 1889; UK Parliament, 1899) imposed a legal requirement for householders and/or general practitioners to notify the sanitary authorities of suspected or diagnosed cases of specific diseases. At the national level, cerebrospinal fever was not a notifiable disease, although circulars issued in autumn 1905 by the Local Government Boards of England and Scotland had suggested, following the recent outbreaks in New York and elsewhere, that the Local Authority should be notified of any suspected outbreaks of meningitis/cerebrospinal fever (Chalmers, 1906, p76). In accordance with these circulars, the Medical O”cer of Health in Glasgow indeed issued a memorandum in May 1906 identifying a possible outbreak (Chalmers, 1906, p76). At subnational level, a clause in the Infectious Disease (Notification) Act 1889 allowed local authorities to use the national notification system to cover additional diseases within their own district by passing a local resolution or order, usually confirmed by the Local Government Board. Invoking this, cerebrospinal fever was made notifiable in Glasgow on 10 August 1906 (Chalmers, 1906, p76). It was initially made notifiable for one year, with this being extended for a further year in 1907, and was added permanently to the list of notifiable diseases in Glasgow in 1908 (Chalmers, 1907, p72). Before 1906, there are no recorded deaths from cerebrospinal fever in Glasgow, although the Medical O”cer of health at the time, Dr Chalmers, notes that a small number may have been mistakenly diagnosed as ‘ordinary’ or ‘tubercular’ meningitis (Chalmers, 1906, p79-80).

The epidemic, thought to have been seeded from that in New York (Osler, 1907), started during 1906. Initially, cases were relatively sporadic. From the second week of January 1907, the epidemic entered what the Medical O”cer of Health referred to as the second or ‘epidemic phase’, with a ‘considerable increase’ in case numbers (Chalmers, 1906, p82, 84), reaching around 30 cases per week. By March, the epidemic was causing as many as 50 cases and 30 deaths per week. Yet, by the end of the first six months of 1907, the epidemic phase was ‘almost over’, with the ‘the number of cases registered during the period January 1st to June 30th being 802, against 196 for the period July 1st to December 31st’ (Chalmers, 1907, p72). The inset of Figure 1A shows the epidemic curve as weekly numbers of cases and deaths from cerebrospinal fever in Glasgow, revealing the dramatic increase during the first half of 1907, followed by a similarly rapid decline by the end of the year. The main panel of Figure 1A, based on annual data, shows the epidemic as a massive spike in 1907, reaching nearly 1,000 cases and 685 deaths in that year, and nearly 1,000 deaths over the period 1906-1908. It also shows a later resurgence in 1915. Figure 1B shows the spatial distribution of mortality rates in 1907, demonstrating that the disease had spread to all areas, but with the highest mortality rates observed in the central and south-eastern wards of the city. Comparing the two left-hand panels of Figure A.2 reveals a similar spatial pattern for incidence rates.

**Figure 1.**
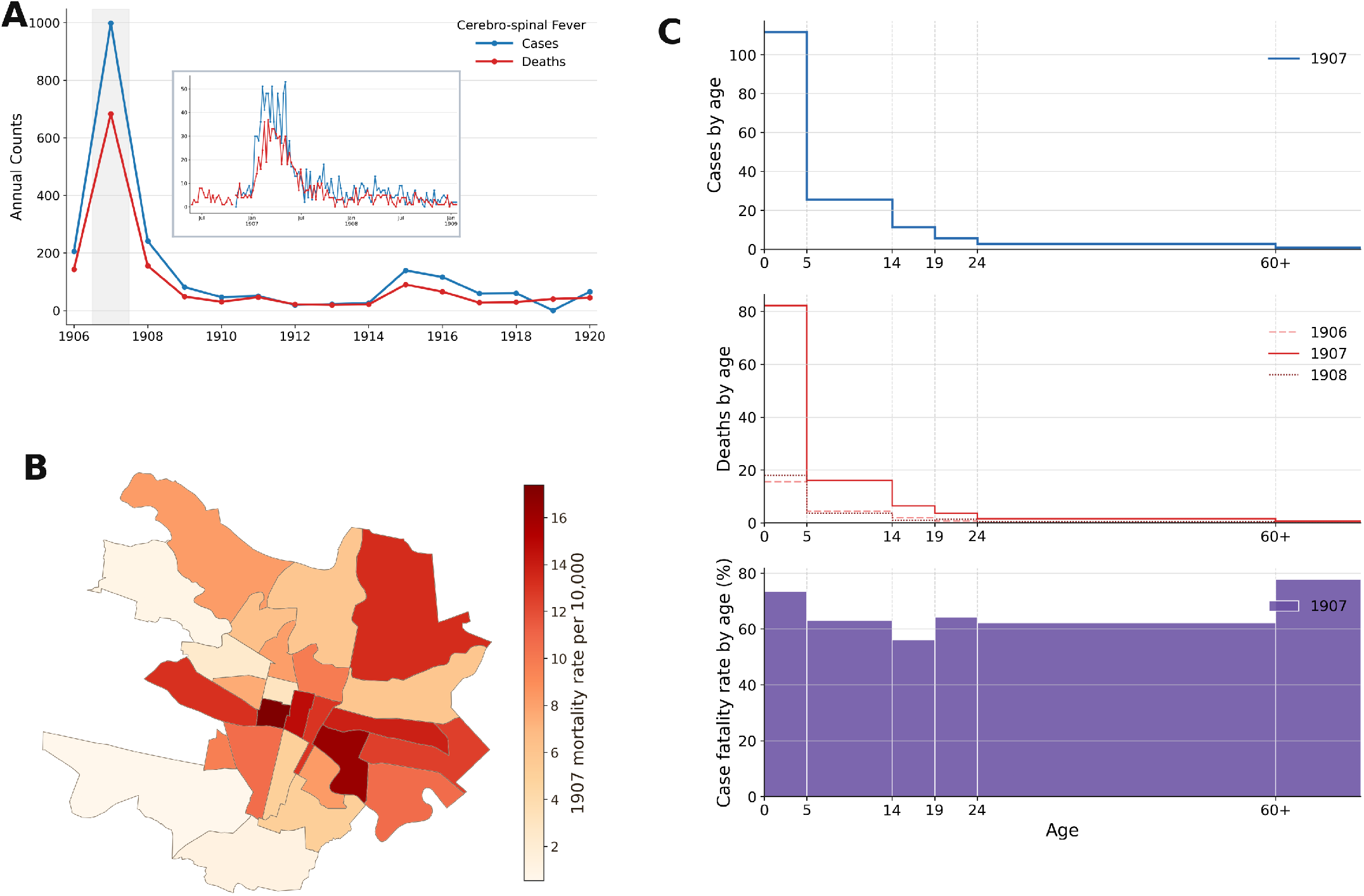
Empirical observations regarding cerebrospinal fever in Glasgow 1906-1920. A: Time series of cases and deaths, 1906-1920; grey shading indicates 1907. A inset: Weekly cases and deaths from cerebrospinal fever over the course of the epidemic. B: Spatial distribution of mortality rate per 10,000 persons by administrative ward of Glasgow in 1907. C: Age distribution of cases (1907), deaths (1906-1908) and case fatality rate (1907).

The epidemic primarily affected children under five, and showed unusually high case fatality rates across all age groups. The top two plots of Figure 1C show the age distribution of cases (blue) and deaths (red) by age group, across the years, revealing the extent to which cases and deaths were seen most strongly in those under the age of five. Figure A.1 presents the same figure by the share of deaths in each age group, demonstrating that the age distribution of deaths differed little between the years of the epidemic. Overall, the outbreak had a very high fatality rate of diagnosed cases: 70% in 1906 (Chalmers, 1906), 68% in 1907 (Chalmers, 1907) and 65% in 1908 (Chalmers, 1908). These rates were high not only relative to local case fatality rates for other infectious diseases – around 10% for smallpox, 8% for whooping cough and 4% for measles and scarlet fever – but su”ciently high even relative to other forms of meningitis, that the Medical O”cer of Health commented that ‘we are dealing with a peculiarly fatal form of meningitis’ (Chalmers, 1906, p82). The bottom plot of Figure 1C reveals that this exceptionally high case fatality rate varied relatively little by age, with only slightly higher case fatality rates among the youngest and oldest age groups. Data analysed by the Medical O”cers (Chalmers, 1907, p76) also showed that deaths often occurred very rapidly, with one third of occurring within the first 2 days. Furthermore, among those who recovered, the illness was often long-lasting, with 30% being ill for over 3 months.

The epidemic caused major concern both locally and further afield. Professor William Osler, writing in the Edinburgh Medical Journal in 1907, noted that ‘At present the disease has broken out in Belfast and Glasgow, and is causing no little alarm’ (Osler, 1907, p200). On 6th February 1907, The Scotsman reported that ‘The Medical O”cer for Glasgow decided to issue daily reports regarding the outbreak’ (Scotsman, 1907, p10). These reports were frequently covered in the newspapers, e.g. in the Scotsman on 7th, 9th, 13th, 14th and 25th of February and 1st and 9th of March. The addition of the disease to the list of notifiable diseases in Glasgow in August 1906 was also reflective of this concern. Treatment during the epidemic was rudimentary, creating further concern. At the time, there was no agreed treatment for cerebrospinal fever. During the epidemic, Drs Currie and McGregor at the local Belvidere Hospital experimented with serum treatment, i.e. injecting humans suffering from cerebrospinal fever with an anti-serum developed in animals. A detailed analysis in the reports concludes that this did not appear to have a marked improvement on survival rates, although it appeared that ‘serum-treated cases which survived the first ten days of illness had a better chance of life’ (Chalmers, 1907, p77). The reports concluded that it would be better to give the serum prophylactically, i.e. before illness becomes established (ibid, p78), but no attempt to do this in practice seems to have been made.

Beyond treatment, although the disease-causing organism was identified, limiting its spread was hampered by knowledge limitations relating to transmission. Biological testing established that the epidemic in Glasgow was of bacterial origin. The microorganism in question, at the time known as *diplococcus intracellularis* and now called *Neisseria meningitidis*, had first been identified in the United States in 1887 (Flexner and Jobling, 1908, p141). Chalmers, the MOH in Glasgow during 1907, wrote that ‘the organism which transmits the infection has been recovered from areas common to the nose and throat’ (Chalmers, 1930, p370). The mode by which the disease was spread, however, was not known. Osler (1907, p200), writing at the time, was aware that unlike other infectious diseases, outbreaks of meningococcal meningitis ‘appear in very widely separated areas, in which it prevails severely, but does not spread widely’, noting that ‘the disease is never pandemic, like influenza, sweeping rapidly over many countries’. An investigation of cases within close proximity conducted early in the epidemic and reported in the annual reports revealed that ‘multiple cases had occurred in 9 houses in which there were altogether 19 cases, there being three in one house […] on the other hand, only one tenement had more than one house infected […] with regard to infectivity, therefore, the risk of direct transference from person to person does not appear to be excessive, but it is obviously much greater to the other members of the invaded household than to other houses in the tenement’ (Chalmers, 1906, p83). Further analysis made it ‘impossible to suggest that the disease was invariably selecting the dirtiest houses; but when one discarded the general term “dirtiness” for one of pressure on air space, the question took a different complexion’ (Chalmers, 1906, p84), which suggested that one of the strongest contributors to cerebrospinal infection and death is what he calls ‘impure air’ (Chalmers, 1906, p83). During the epidemic phase, ‘multiple cases in families also became more numerous and with this came a suggestion that the essential conditions of spread lay in domestic overcrowding’ (Chalmers, 1930, p371). The analysis also excluded a strong role for transmission within schools (Chalmers, 1906, p88-9), and for professional occupation (Chalmers, 1906, p89), but ‘place groupings without any other association are suggested’ (Chalmers, 1906, p90). Chalmers concluded that the most likely means of transmission is when the weather ‘promotes catarrhal conditions’ which encouraged people to stay indoors, leading to ‘pressure on air space’ (Chalmers, 1906, p84). Interestingly, Chalmers posited that the infection could be carried by healthy individuals so that the disease was reintroduced to families even after the house had been thoroughly disinfected (Chalmers, 1930, p372). Indeed, he notes that ‘the organism was recovered… from the naso-pharynx of 4 [asymptomatic] contacts, and it is suggestive that those were inmates of the houses in which 4 and 5 cases respectively had occurred’ (Chalmers, 1906, p90). The presence of asymptomatic carriers was subsequently shown to be the case but was still a supposition during Glasgow in the 1907 epidemic. In summary, Chalmers concluded that “‘carrier” cases and spray-infection would completely explain the method of spread were the organism recoverable in a larger proportion of contacts’ (Chalmers, 1906, p90). The importance of close contact was also consistent with the observation that although a similar epidemic, although smaller in scale, occurred in Belfast (Robb, 1907), and a small number of cases were observed in Liverpool, the disease remained relatively confined to Glasgow.

In summary, the Medical O”cers considered that transmission was driven by close contact, often with multiple cases in the same household, and affected by the space available within an accommodation; although cases clustered in space, there was apparently limited impact of between-household proximity. General markers of poverty and ‘dirt’ were not apparently associated with transmission, and, despite the fact that children were disproportionately affected, schools did not seem to be a major contributor to transmission. Nevertheless, each of these factors was analysed separately in the Medical O”cer report. We therefore conduct analysis to assess the extent to which the same drivers are observed when considering them jointly, and when controlling for socioeconomic conditions and population structure.

## 3 Determinants of incidence and mortality

In our analysis, we first exploit the variation in incidence and mortality rates across the administrative units of Glasgow, namely municipal wards, to assess the contribution to incidence and mortality rates of the factors raised by the Medical O”cers. We estimate statistical models of incidence and mortality rates on proxies of: close contact based on the availability of space within accommodation (persons per house and windowed rooms per house), and housing density (inhabited houses per square kilometre). In these regressions, we control for markers of population structure (birth rate per 1,000 population) and poverty (percentage of illegitimate births), and allow unobservable factors to be correlated between wards when computing standard errors. Alternative model specifications are discussed in Methods and we report robustness to these specifications in Supplementary Materials.

Figure 2 shows model coe”cient estimates and 95% confidence intervals from linear regressions, for incidence rate in blue and mortality rate in red. The first observation is that the pattern is very similar between incidence and mortality rates, reflecting the high correlation between these two variables. Full results shown in Table A.3 include *R*^2^ values showing that the model explains over 65% of the variation in incidence and mortality rates across administrative wards. Regarding markers of close contact, the estimates reveal that persons per house is positively related with incidence and mortality rates, while windowed rooms per house is negatively related, consistent with heightened within-house transmission in houses with more individuals and less space available. A one standard deviation increase in the number of persons per house (i.e. an increase of 0.27 persons; see Table A.2 for descriptive statistics for 1907) increased the incidence rate by 2.9 per 10,000 persons, while a 1 standard deviation decrease in windowed rooms (i.e. a decrease of 1.3 rooms), increased it by 4.4. Regarding housing density, the effect of number of inhabited houses per square kilometre is estimated to be near zero and not statistically significantly different from zero, for both incidence and mortality rates, consistent with the pattern of limited between-house transmission observed by the Medical O”cers. Regarding population structure, we control for the birth rate per 1,000 population to account for potential confounding due to differing proportions of infants between wards, given the high incidence among this age group. The coe”cient is not significant, although tending to be positive, informing us that the pattern of cases was not strongly driven by the proportion of infants in a ward, and confirms that close contact was important, over and above any effect from population structure. Regarding our marker of poverty, the percentage of births that were illegitimate, this variable is positively related with incidence rates and close to significance for mortality. Although this marker of poverty potentially captures many drivers, it is also consistent with the poorest individuals living in the closest contact with others. The residuals from these regressions, shown spatially on a map of Glasgow in the right-hand panel of Figure A.2, suggest a spatial pattern. In general, the findings are consistent with the analysis of the Medical O”cers, suggesting that the size of houses and the number of people living in them were key drivers of risk.

**Figure 2.**
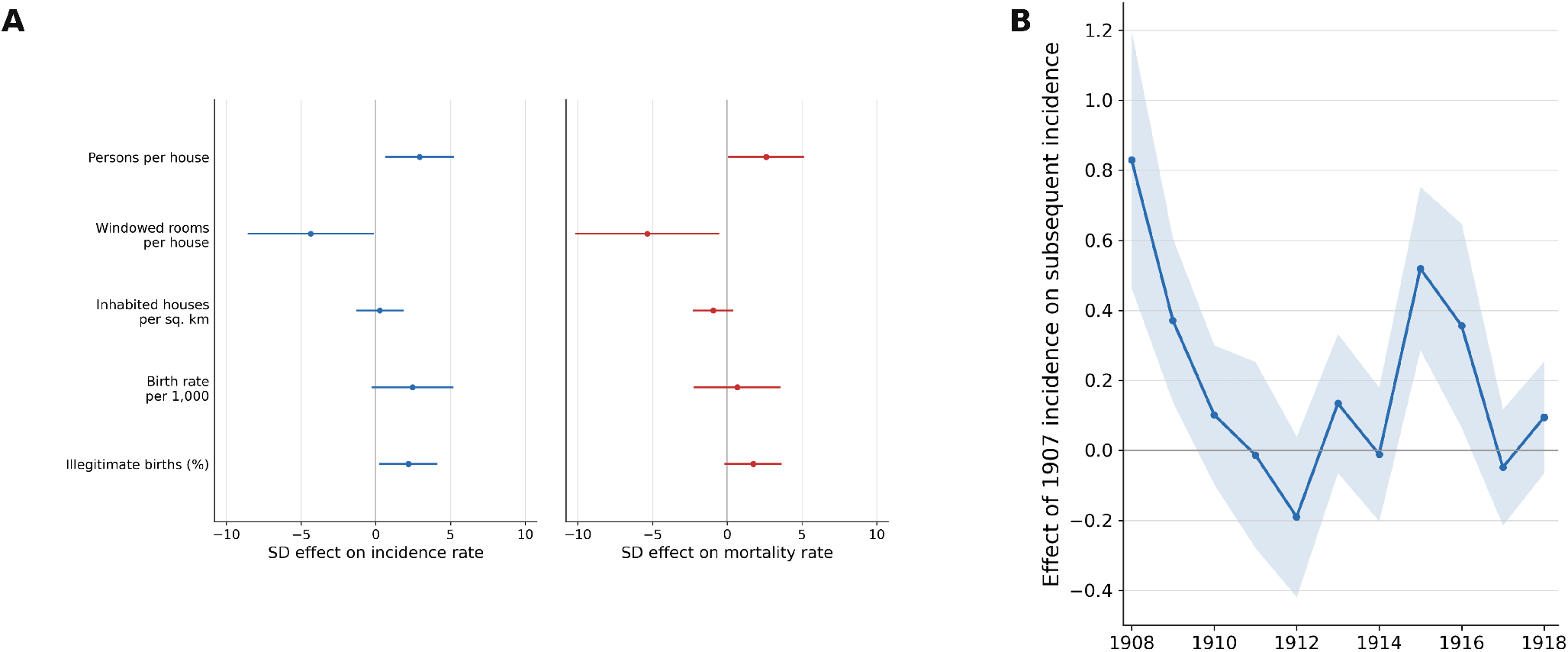
Baseline model results on the drivers of 1907 incidence and mortality rates and on the effect of 1907 incidence rates on subsequent incidence rates. Panel A shows 1 SD effects and 95% confidence intervals from ward-level linear models for 1907 incidence and mortality rates per 10,000 population. Panel B shows year-specific 1 SD effects of the 1907 incidence rate on subsequent incidence rates from pooled ward-year linear models with year fixed effects and interactions between 1907 incidence rates and year indicators; shaded areas denote 95% confidence intervals.

We conducted robustness testing to alternative model specifications and control variables. In addition to the linear model, we also estimated a Poisson model based on counts of cases and deaths (Table A.3), controlling for population size in the ward. In this specification, although persons per house was no longer significant, windowed rooms per house gained in significance, as did the percentage of illegitimate births. In this model, inhabited houses now appears to have a slightly negative effect on mortality rates. Other variables retained their significance and direction (see Figure A.3).

In our baseline results, we had used a 2 kilometre cut-off for ward centroid distances, which ensures that no ward is left spatially isolated (each ward has at least one neighbour). The centroid distance between the most isolated ward (Springburn) and the closest neighbour is 1.8km. Overall, the minimum distance between ward centroids is 0.46km, with a median of 2.98km. We also conducted robustness using the linear model to using different spatial cut-offs (1.5km and 2.5km) (Table A.4), finding similar significance. Finally, we replaced the birth rate per 1,000 population in 1907 with the proportion of the population aged under than 5 years in 1901 (Chalmers, 1902b). This variable was not significant and this change had little effect on the other variables.

Second, we then examined the effect of incidence rates in 1907 on incidence rates in subsequent years. To do this, we employed a panel specification as described in Methods, controlling for the same observables as those in Figure 2A. The results are shown in Figure 2B, with full results of the estimated model shown in Table A.6. We find that higher incidence rates in a ward during the 1907 epidemic year are related to higher risk for the following 2-3 years, over and above effects of the observables controlled for in panel A, which themselves are not significant predictors; a similar effect is seen during the later outbreak in 1915. That cases continued at higher frequency in the same areas into 1908 and 1909 can be explained by the presence of ongoing transmission and carrier cases. However, that the effect reappears in 1915 suggests that there are persistent underlying factors that make populations in particular areas more vulnerable. This wartime outbreak is thought to have arisen as a local manifestation of a general increase in prevalence driven primarily by close contact and fatigue among soldiers housed in poorly ventilated barracks (Peters and Gunn, 1930); there are also reports of cases onboard ships arriving in Glasgow (Chalmers, 1919).

## 4 Discussion

Relative to meningococcal meningitis outbreaks in temperate regions today, the epidemic in Glasgow in 1906-1908 was very large, with a much higher case fatality rate. Overall, cases of meningococcal meningitis have been falling over the last few decades in the UK (Figure A.4), in part likely due to the introduction of meningitis vaccination programmes. Today, case fatality rates from meningococcal meningitis are estimated to be around 10% (Public Health Wales, 2026; National Health Service, 2026), with only around 5% of cases occurring in clusters (Public Health Wales, 2026). Recent outbreaks in the UK include two cases seen in a nursery in 2023 (Mahase, 2026) or 65 cases between October 1981 and March 1986 in Gloucestershire (Cartwright et al., 1986). The 2026 outbreak in Kent, UK involves 21 confirmed cases during March 2026 (UK Health Security Agency, 2026) and two deaths. All of these are several orders of magnitude smaller than the epidemic in Glasgow in 1907.

Historical experience helps reveal that even though a very large proportion of the variation in incidence between administrative areas during the 1907 epidemic was explained by observable factors, in studying subsequent outbreaks, unobserved factors captured by prior incidence rates in 1907 contributed to predicting incidence rates. From the perspective of the basic science of epidemiology, this persistence suggests that the emergence of meningococcal clusters may not occur randomly in space, perhaps encouraging investigations to attempt to understand why populations in some locations are more at risk than others. From a public health perspective, persistence suggests that in the absence of precise drivers of incidence, it makes sense to use prior exposure of a population as a proxy for disease risk. This, for example, highlights the value of spatial information for prioritising antibiotic prophylaxis and vaccination. Indeed, our findings would suggest that roll-out of such interventions might reasonably start with populations in areas that have historically experienced higher incidence.

## 5 Conclusion

Using Medical O”cer of Health records for Glasgow and other archival sources, we analysed incidence and mortality rates from the largest epidemic of meningitis on record in the British Isles, which peaked in 1907. We examined the time evolution of cases and deaths and their spatial pattern across the municipal areas of the city, as well as their distribution over age groups, reviewing knowledge at the time and public health responses. We then exploited the variation in cases and mortality between municipal areas to assess the role of contributing factors, enhancing the analysis conducted at the time by controlling for socioeconomic factors. We found that within-household close contact, as captured by house size, was the most robust correlate of both incidence and mortality rates. We then examined the persistence of the patterns of disease incidence rates during the fifteen years after the main epidemic wave. We found that populations in areas that showed the highest incidence rates in 1907 remained at higher risk for the following two years (even after controlling for observable factors), and were also more strongly affected in a subsequent outbreak in 1915.

Our findings have implications in the context of the recent meningitis outbreak in Kent, UK, and for the management and mitigation of meningococcal meningitis clusters in temperate regions going forward. Firstly, they show that outbreaks of bacterial meningitis of this scale or explosiveness are far from unprecedented in the UK. Secondly, they confirm the importance of overcrowding and close contact for transmission. Thirdly, they highlight the importance of spatially localised and targeted antibiotic delivery and vaccination in affected areas to avoid recurrent cases in affected areas over the coming years. Fourthly, the persistence of location-specific but unidentified risk factors nearly a decade after the main epidemic is suggestive of the potential to uncover currently unknown but persistent drivers of risk.

## 6 Methods

### Data

During the period of interest, the city of Glasgow was divided into 26 geographical administrative areas, called municipal wards, which are used in the analysis. The city was divided into these areas to facilitate the understanding of the impact of differing local conditions on health, or, in the words of the then Medical O”cer of Health, to allow ‘[v]ital statistics … [to be] viewed in the light of an intimate knowledge of the local circumstances which influence them’ (Chalmers, 1902a). Data on the cerebrospinal fever outbreak in Glasgow, 1906-1908 were sourced from the Medical O”cer of Health (MOH) Reports for Glasgow 1906, 1907 and 1908 (Chalmers, 1906, 1907, 1908). The reports provided numbers for cases and deaths for the city as a whole, and cases and deaths for the individual wards of the city from 1906 to 1920, initially in the MOH reports (Chalmers, 1906, 1907, 1908, 1909, 1910, 1911, 1912, 1913) and then in loose-leaf papers held in the Glasgow City Archives. Data on cases and deaths of cerebrospinal fever by age groups at the city and ward level were also available for 1907. Weekly data on cases and deaths of cerebrospinal fever (at the level of the city) were sourced from tables in the Glasgow Medical Journal. Population size was also provided by the MOH reports at ward level. Some socioeconomic markers for the wards were provided directly in the MOH reports (birth rate, percentage of illegitimate births) and others were calculated from the counts of population and number of inhabited houses, both of which were available annually in the MOH reports. The Under-5 share of the population was provided in the Report on the 1901 Census Chalmers (1902b). According to usage at the time, the term ‘house’ referred to an individual dwelling. In Glasgow, most people lived in flats in tenement blocks, although individual houses were common in areas further from the city centre; for most households, the notion of a ‘house’ therefore corresponds to a flat. The number of inhabited houses per square kilometre was calculated from the number of inhabited houses and the acreage of the ward, reported in the MOH reports. Persons per windowed room values were only provided for 1901 (Chalmers, 1902a) and 1911 (Chalmers, 1912) and are interpolated for intervening years. These then permitted the calculation of windowed rooms per house. Data were transcribed from the various sources into an Excel spreadsheet to permit analysis.

We use polygon shapefiles of the municipal ward boundaries created by Mancy (2023) to represent the spatial dimension of the data. These are used in Figure 1 and Figure A.2. We additionally use these shapefiles to assess spatial correlation in the 1907 outbreak outcomes. Specifically, we calculate Moran’s *I* for ward-level incidence and mortality rates in 1907 using queen-contiguity weights. We find positive spatial correlation in incidence, with Moran’s *I* = 0.198 and a two-sided permutation *p*-value of 0.049, and slightly weaker evidence for mortality, with Moran’s *I* = 0.167 and a two-sided permutation *p*-value of 0.089. Finally, we use the shapefiles to compute ward centroids, which are used to adjust the standard errors in the linear models described below for spatial correlation.

### Statistical Models

We assess the determinants of outbreak cases and mortality in 1907 using a linear specification in which the outcomes – ward-level incidence and mortality rates – are expressed as a function of observable ward-level characteristics and unobservable idiosyncratic factors. In particular, for our baseline specification, we estimate

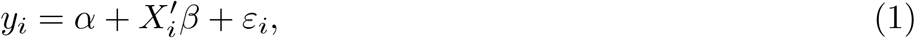

for ward *i* in 1907, where *y*_*i*_ is either the 1907 incidence rate per 10,000 or the 1907 mortality rate per 10,000, and *X*_*i*_ contains ward characteristics: *Persons per house, Windowed rooms per house, Inhabited houses per sq. km, Birth rate per 1,000, Illegitimate births (%)*. Descriptive statistics of the variables included in our statistical analysis for all available years are in Table A.1; those for 1907 are provided in Table A.2. We report results from least squares estimation of equation (1) in Figure 2 (panel A) in the main text and Table A.3 in Supplementary Material.

We also estimate a Poisson model for cases and deaths accounting for population size:

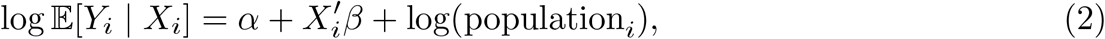

where *Y*_*i*_ denotes the number of cases or deaths. Results are reported in Table A.3 in Supplementary Material and in Figure A.3.

Unobserved determinants of epidemic severity in the error term *ε*_*i*_ may themselves be spatially clustered across wards. Indeed, Supplementary Figure A.2C and D map the residuals from the 1907 cross-sectional regressions for incidence and mortality after conditioning on observed characteristics, showing that the residuals remain spatially correlated. To account for spatial dependence, we thus compute Conley spatial heteroskedasticity-and-autocorrelation-consistent standard errors with a Bartlett kernel and a 2 km cutoff. We use these standard errors to calculate confidence intervals and p-values. Our results are robust to alternative Conley cut-off distances of 1.5 km and 2.5 km; see Supplementary Material Table A.4.

Disease incidence and mortality was higher among children. To account for potential variation in demographic composition across wards that may affect remaining factors we study, we have included in our baseline model specification the birth rate (per 1,000 population) of the ward, a proxy for the proportion of the population that are infants. We conducted robustness analysis to the under-5 population share, measured in 1901 (Chalmers, 1902b). The birth rate in 1907 and the under-5 population share (in 1901) are highly correlated (*r* = 0.964, pairwise; *N* = 25). Indeed, using the under-5 population share instead of birth rate in our model does not alter the results. Results of these regressions are reported in Table A.5 in Supplementary Material. The advantage of the birth rate is that it is also available at the level of the ward for the years after 1907, and thus is useful for the analysis of dynamic effects we next turn to, whereas the under-5 population share is only available for 1901.

To examine the persistence of incidence during the main epidemic waves on future incidence rates, we estimate

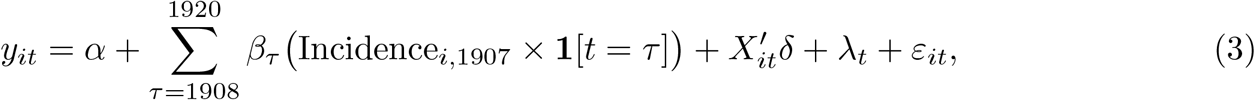

where *y*_*it*_ is the incidence rate in ward *i* and year *t, X*_*it*_ is the time-varying control set used for the model in equation (1), and λ_*t*_ are year fixed effects. This specification allows the effect of the epidemic incidence rate in 1907 on subsequent years to vary flexibly over time. We report the estimated coe”cients *β*_τ_ for each year in Figure 2B in the main text.

## Author contributions

**Gillian Stewart:** Data curation, Investigation, Visualisation, Writing – original draft, Writing – review & editing.

**Max Schroeder:** Data curation, Formal analysis, Methodology, Software, Visualisation, Writing – original draft, Writing – review & editing.

**Rebecca Mancy:** Conceptualisation, Funding acquisition, Methodology, Project administration, Supervision, Visualisation, Writing – original draft, Writing – review & editing

**Konstantinos Angelopoulos:** Conceptualisation, Funding acquisition, Methodology, Project administration, Supervision, Writing – original draft, Writing – review & editing

## Rights retention

For the purpose of open access, the authors have applied a Creative Commons Attribution (CC BY) licence to any Author Accepted Manuscript version arising from this submission.

## Data availability

All data utilised in this study are publicly available, with sources provided in the main text and metadata provided in the files associated with the dataset. The dataset analysed here is available with the code for the paper.

## Code availability

The code repository for the study is available at: https://doi.org/10.5281/zenodo.19589744.

## Bibliography

Cartwright, K. V., Stuart, J., and Noah, N. (1986). An outbreak of meningococcal disease in gloucestershire. The Lancet, 328(8506):558–561.

Chalmers, A. (1902a). Report of the medical officer of health of the city of glasgow. Glasgow, UK: Corporation of Glasgow. See https://wellcomecollection.org/works/e7sa7xz7.

Chalmers, A. (1905). Report of the medical officer of health of the city of glasgow. Glasgow, UK: Corporation of Glasgow. See https://wellcomecollection.org/works/b6jpbw7c.

Chalmers, A. (1906). Report of the medical officer of health of the city of glasgow. Glasgow, UK: Corporation of Glasgow. See https://wellcomecollection.org/works/vm6m8gjj.

Chalmers, A. (1907). Report of the medical officer of health of the city of glasgow. Glasgow, UK: Corporation of Glasgow. See https://wellcomecollection.org/works/dcrma5qe.

Chalmers, A. (1908). Report of the medical officer of health of the city of glasgow. Glasgow, UK: Corporation of Glasgow. See https://wellcomecollection.org/works/jmshfe9d.

Chalmers, A. (1909). Report of the medical officer of health of the city of glasgow. Glasgow, UK: Corporation of Glasgow. See https://wellcomecollection.org/works/zrvhbem6.

Chalmers, A. (1910). Report of the medical officer of health of the city of glasgow. Glasgow, UK: Corporation of Glasgow. See https://wellcomecollection.org/works/gxhbuj6m.

Chalmers, A. (1911). Report of the medical officer of health of the city of glasgow. Glasgow, UK: Corporation of Glasgow. See https://wellcomecollection.org/works/drdbhxu9.

Chalmers, A. (1912). Report of the medical officer of health of the city of glasgow. Glasgow, UK: Corporation of Glasgow. See https://wellcomecollection.org/works/ky4ak4j4.

Chalmers, A. (1913). Report of the medical officer of health of the city of glasgow. Glasgow, UK: Corporation of Glasgow. See https://wellcomecollection.org/works/m6sf8cgs.

Chalmers, A. (1919). Report of the medical officer of health of the city of glasgow, 1914–1918. Glasgow, UK: Corporation of Glasgow. https://wellcomecollection.org/works/rjbwmyfg.

Chalmers, A. K. (1902b). Census 1901-Report on Glasgow Its Sanitary Districts and Municipal Wards. Corporation of Glasgow.

Chalmers, A. K. (1930). The health of Glasgow, 1818-1925: an outline. authority of the Corporation.

Chang, Q., Tzeng, Y.-L., and Stephens, D. S. (2012). Meningococcal disease: changes in epidemiology and prevention. Clinical epidemiology, pages 237–245.

Flexner, S. and Jobling, J. W. (1908). Serum treatment of epidemic cerebro-spinal meningitis. The Journal of experimental medicine, 10(1):141.

Mahase, E. (2026). Meningitis: Menb strain may have evolved to be more transmissible, say experts. BMJ, 392.

Mancy, R. (2023). Shapefiles of administrative boundaries, subway and main rivers in glasgow, uk, around 1910. Zenodo.

National Health Service (2026). Meningitis. Accessed 23 April 2026.

Osler, W. (1907). Cerebro-spinal fever. Edinburgh Medical Journal, 21(3):199.

Parliament of the United Kingdom (1889). Infectious disease (notification) act 1889. UK Public General Act, 52 & 53 Vict. c. 72; accessed 23 April 2026.

Peters, R. and Gunn, W. (1930). Cerebro-spinal fever in glasgow, 1929. Epidemiology & Infection, 30(4):420–432.

Public Health Wales (2026). Meningitis and meningococcal disease. Accessed 23 April 2026.

Robb, A. G. (1907). Some observations on the recent outbreak of cerebro-spinal fever in belfast. The British Medical Journal, pages 1129–1131.

Schroeder, M., Lazarakis, S., Mancy, R., and Angelopoulos, K. (2023). An Extended Period of Elevated Influenza Mortality Risk Follows the Main Waves of Influenza Pandemics. Social Science & Medicine, 328:115975.

Scotsman (1907). The cerebrospinal fever outbreaks. Scotsman.

Smith, F. (1995). The russian influenza in the united kingdom, 1889–1894. Social History of Medicine, 8(1):55–73.

UK Health Security Agency (2025). Laboratory confirmed cases of invasive meningococcal infection in england: annual report 2024 to 2025 data tables. Supplementary data workbook (Excel), Table 2.

UK Health Security Agency (2026). Notified cases of invasive meningococcal disease. Accessed 23 April 2026.

UK Parliament (1899). Infectious disease (notification) act (1889) extension bill. House of Commons debate, 3 March 1899; accessed 23 April 2026.

