## Supplementary Materials for "Spatial variation in incidence of meningococcal meningitis: evidence from a large historical epidemic in Glasgow"

### A Supplementary Materials

#### A.1 Additional Tables and Figures

Table A.1: Summary Statistics 1906-1920, all years combined

| Variable | N | Mean | SD | Min | Max |
|---|---|---|---|---|---|
| Population | 390 | 30601 | 13573 | 1442 | 57211 |
| Cases | 364 | 5.541 | 11.764 | 0.000 | 80.000 |
| Deaths | 375 | 3.813 | 8.310 | 0.000 | 60.000 |
| Incidence rate per 10,000 | 364 | 1.804 | 3.567 | 0.000 | 22.247 |
| Mortality rate per 10,000 | 375 | 1.191 | 2.535 | 0.000 | 17.375 |
| Persons per house | 390 | 4.735 | 0.263 | 4.111 | 5.471 |
| Inhabited houses per sq. km | 390 | 4817 | 3079 | 583 | 12956 |
| Windowed rooms per house | 364 | 2.756 | 1.157 | 1.575 | 6.643 |
| Birth rate per 1,000 | 390 | 25.57 | 8.48 | 6.37 | 41.70 |
| Under-5 share of population (1901) | 25 | 0.112 | 0.031 | 0.051 | 0.149 |
| Illegitimate births (%) | 390 | 8.41 | 5.52 | 2.06 | 45.46 |

*Notes:* Number of observations varies because some source variables are unavailable in specific ward-years. The analysis sample contains 390 ward-year observations (26 wards x 15 years) after dropping administrative area referred to as 'Institutions and Harbour'. Cases and incidence are unavailable for all wards in 1919; deaths and mortality are unavailable for 15 wards in 1912; windowed rooms per house is unavailable for all wards in 1920; and the under-5 share is unavailable for Kinning Park.

Table A.2: Summary Statistics 1907

| Variable | N | Mean | SD | Min | Max |
|---|---|---|---|---|---|
| Population | 26 | 30169 | 12952 | 2035 | 49577 |
| Cases | 26 | 37.231 | 24.916 | 3.000 | 80.000 |
| Deaths | 26 | 25.962 | 18.509 | 1.000 | 60.000 |
| Incidence rate per 10,000 | 26 | 12.353 | 6.096 | 1.337 | 22.247 |
| Mortality rate per 10,000 | 26 | 8.625 | 4.825 | 0.540 | 17.375 |
| Persons per house | 26 | 4.806 | 0.272 | 4.478 | 5.466 |
| Inhabited houses per sq. km | 26 | 4653 | 2959 | 658 | 10794 |
| Windowed rooms per house | 26 | 2.843 | 1.343 | 1.620 | 6.357 |
| Birth rate per 1,000 | 26 | 28.023 | 9.500 | 8.203 | 39.888 |
| Under-5 share of population (1901) | 25 | 0.112 | 0.031 | 0.051 | 0.149 |
| Illegitimate births (%) | 26 | 7.606 | 5.085 | 2.632 | 26.316 |

*Note:* Summary statistics use the 1907 cross-section of the 26-ward analysis sample after dropping administrative area referred to as 'Institutions and Harbour'. Under-5 share of population is unavailable for Kinning Park.

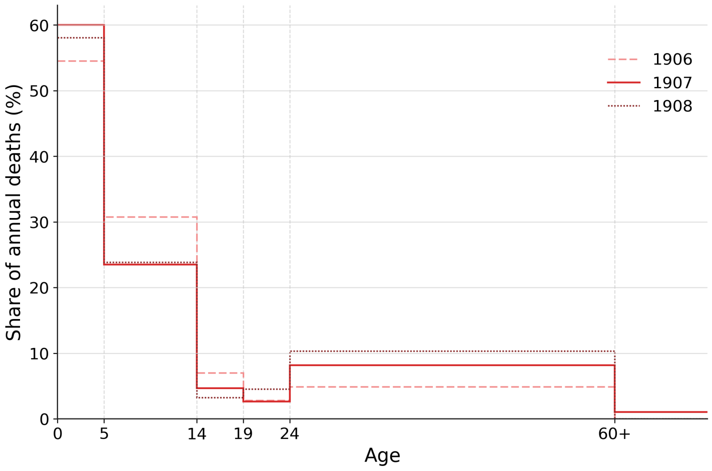


Figure A.1: **Age distribution of deaths (1906-1908), presented as shares of the deaths during each year 1906-1908.**

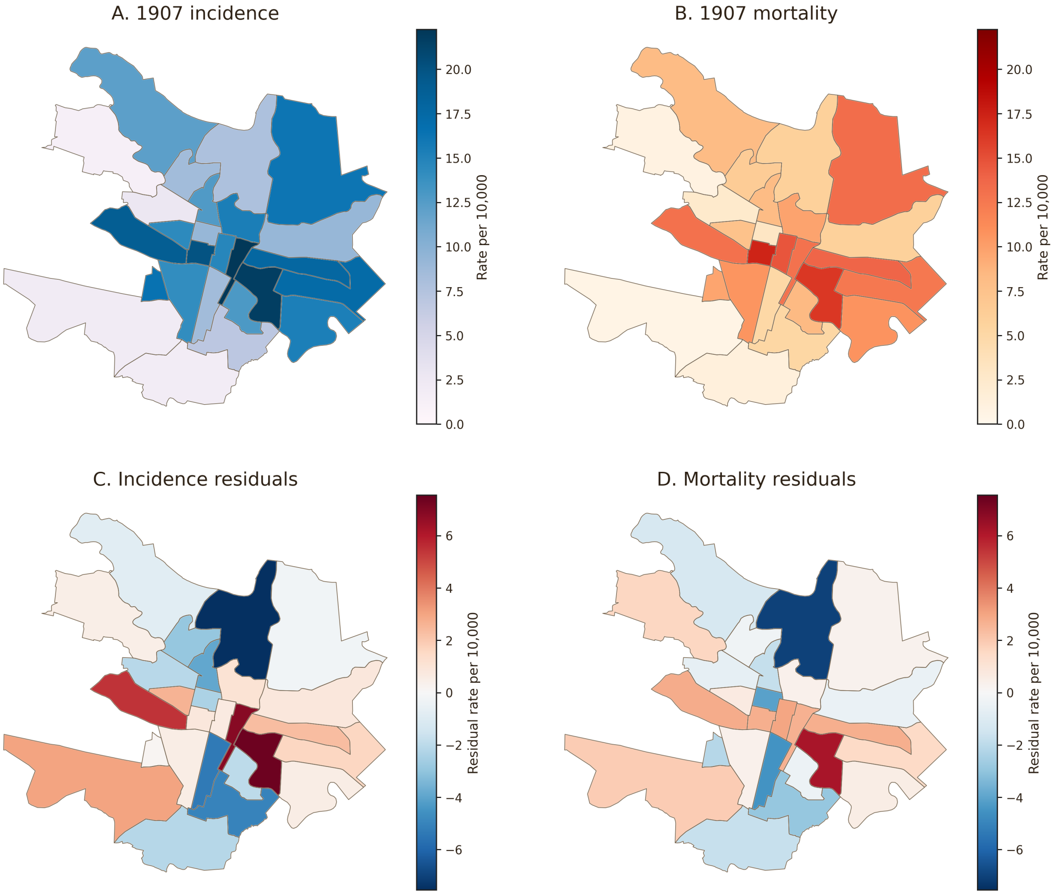


Figure A.2: **Spatial distribution of epidemic incidence, mortality, and regression residuals across Glasgow wards in 1907.** Panels A and B map the incidence and mortality rates per 10,000 population. Panels C and D map residual incidence and residual mortality from the corresponding 1907 cross-sectional regressions after conditioning on observed ward characteristics.

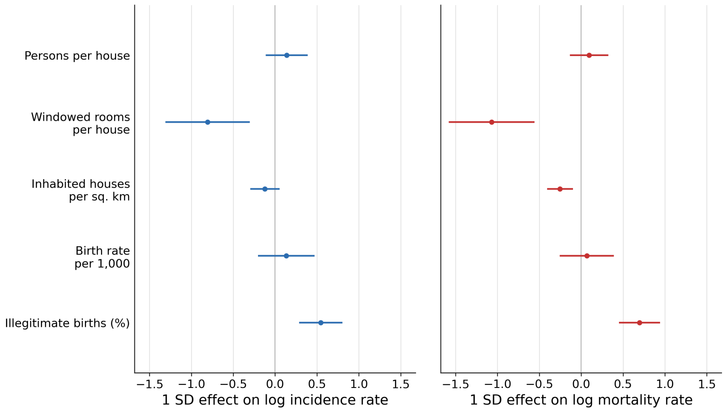


Figure A.3: **Poisson model results on the drivers of 1907 incidence and mortality rates.** The figure shows 1 SD effects and 95% confidence intervals from ward-level Poisson models for 1907 incidence and mortality rates per 10,000 population.

Table A.3: Linear and Poisson Estimates of 1907 Incidence and Mortality

| | Linear models | | Poisson models | |
|---|---|---|---|---|
| | Incidence (1) | Mortality (2) | Cases (3) | Deaths (4) |
| Persons per house | 10.787 | 9.588 | 0.512 | 0.347 |
| | [0.012] | [0.044] | [0.278] | [0.418] |
| Windowed rooms per house | -3.240 | -3.986 | -0.601 | -0.796 |
| | [0.044] | [0.030] | [0.002] | [0.000] |
| Inhabited houses per sq. km | 0.000 | -0.000 | -0.000 | -0.000 |
| | [0.739] | [0.177] | [0.174] | [0.001] |
| Birth rate per 1,000 | 0.259 | 0.071 | 0.014 | 0.007 |
| | [0.078] | [0.651] | [0.442] | [0.681] |
| Illegitimate births (%) | 0.428 | 0.343 | 0.107 | 0.137 |
| | [0.029] | [0.074] | [0.000] | [0.000] |
| Observations | 26 | 26 | 26 | 26 |
| R-squared | 0.66 | 0.66 | | |
| Log likelihood | | | -99.55 | -79.97 |

*Notes:* Columns (1)–(2) report linear regressions of ward-level incidence and mortality rates in 1907, estimated with Conley (1999) spatial HAC standard errors using a Bartlett kernel and a 2 km cutoff using ward-centroid distance. Columns (3)–(4) report Poisson quasi-maximum-likelihood estimates for counts of cases and deaths, including ward population as an exposure term and using heteroskedasticity-robust standard errors. Coefficients in columns (3)–(4) are interpreted as log-rate effects. Bracketed entries report p-values.

Table A.4: Robustness of Cross-Sectional Estimates to Spatial HAC Cutoff Distance

| | 1.5 km cutoff | | 2.5 km cutoff | |
|---|---|---|---|---|
| | (1) | (2) | (3) | (4) |
| Persons per house | 10.787 | 9.588 | 10.787 | 9.588 |
| | [0.019] | [0.052] | [0.008] | [0.025] |
| Windowed rooms per house | -3.240 | -3.986 | -3.240 | -3.986 |
| | [0.053] | [0.029] | [0.038] | [0.026] |
| Inhabited houses per sq. km | 0.000 | -0.000 | 0.000 | -0.000 |
| | [0.777] | [0.169] | [0.703] | [0.173] |
| Birth rate per 1,000 | 0.259 | 0.071 | 0.259 | 0.071 |
| | [0.103] | [0.658] | [0.054] | [0.634] |
| Illegitimate births (%) | 0.428 | 0.343 | 0.428 | 0.343 |
| | [0.023] | [0.060] | [0.025] | [0.054] |
| Observations | 26 | 26 | 26 | 26 |
| R-squared | 0.66 | 0.66 | 0.66 | 0.66 |

*Notes:* This table reports cross-sectional regression estimates for 1907 outcomes under alternative assumptions about the spatial correlation structure of the error term. Standard errors are computed using Conley (1999) spatial HAC estimators with a Bartlett kernel. Columns (1)–(2) are for incidence and mortality rates using a 1.5 km cutoff radius, while columns (3)–(4) are for incidence and mortality rates using a 2.5 km cutoff. Bracketed entries report p-values.

Table A.5: Robustness to Alternative Controls: Age Structure

| | Under-5 share of population | |
|---|---|---|
| | (1) | (2) |
| Persons per house | 10.902 | 10.196 |
| | [0.066] | [0.057] |
| Windowed rooms per house | -4.367 | -5.254 |
| | [0.002] | [0.000] |
| Inhabited houses per sq. km | 0.000 | -0.000 |
| | [0.983] | [0.037] |
| Under-5 share of population | 35.153 | -27.539 |
| | [0.550] | [0.577] |
| Illegitimate births (%) | 0.471 | 0.336 |
| | [0.031] | [0.089] |
| Observations | 25 | 25 |
| R-squared | 0.64 | 0.67 |

*Notes:* This table reports robustness checks for the 1907 cross-sectional regression. Standard errors are computed using Conley (1999) spatial HAC estimators with a Bartlett kernel and a 2 km cutoff. Columns (1)–(2) are for incidence and mortality rates respectively, replacing the birth rate with the share of the population aged under 5. Bracketed entries report p-values.

Table A.6: Effect of 1907 incidence on subsequent incidence 1908–1918

| | Subsequent incidence<br>(1) |
|---|---|
| 1907 incidence effect in 1908 | 0.139 |
| | [0.000] |
| 1907 incidence effect in 1909 | 0.062 |
| | [0.002] |
| 1907 incidence effect in 1910 | 0.017 |
| | [0.316] |
| 1907 incidence effect in 1911 | -0.002 |
| | [0.923] |
| 1907 incidence effect in 1912 | -0.032 |
| | [0.104] |
| 1907 incidence effect in 1913 | 0.022 |
| | [0.185] |
| 1907 incidence effect in 1914 | -0.002 |
| | [0.905] |
| 1907 incidence effect in 1915 | 0.087 |
| | [0.000] |
| 1907 incidence effect in 1916 | 0.059 |
| | [0.017] |
| 1907 incidence effect in 1917 | -0.008 |
| | [0.565] |
| 1907 incidence effect in 1918 | 0.016 |
| | [0.245] |
| Persons per house | 0.590 |
| | [0.131] |
| Windowed rooms per house | -0.480 |
| | [0.096] |
| Inhabited houses per sq. km | -0.000 |
| | [0.121] |
| Birth rate per 1,000 | -0.039 |
| | [0.248] |
| Illegitimate births (%) | 0.005 |
| | [0.791] |
| Observations | 286 |
| R-squared | 0.54 |
| Year fixed effects | Yes |

*Notes:* Column (1) reports the pooled post-1907 ward-year linear model for 1908–1920 estimated with Conley (1999) HAC standard errors, a Bartlett kernel, and a 2 km cut-off. Numbers in square brackets are p-values. Outbreak-effect rows report unscaled year-specific linear combinations. Year fixed effects are included but not displayed.

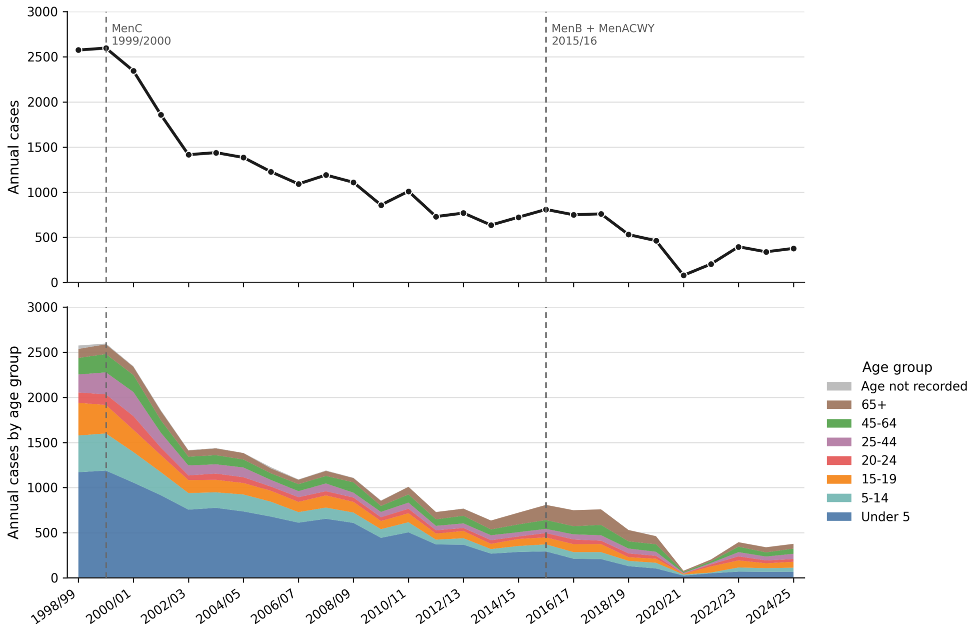


Figure A.4: **Trends in laboratory-confirmed invasive meningococcal disease in England 1998–2025.** The figure plots laboratory-confirmed invasive meningococcal disease (IMD) cases in England by epidemiological year, where each year runs from July to June. The data are from UKHSA's annual supplementary workbook for the 2024/25 IMD report, Table 2, which reports cases by age group from 1998/99 to 2024/25 (UK Health Security Agency, 2025). The lower panel collapses the published age categories as follows: Under 5 combines Under 1 and ages 1 to 4; 5-14 combines ages 5 to 9 and 10 to 14; 65+ corresponds to the published older-age category. In earlier years of the data series, a small 'Age not recorded' category is included where the published total exceeds the sum of the reported age bins. Vertical dashed lines mark the introduction of the MenC vaccination programme in 1999 and the infant MenB plus teenage MenACWY programmes in 2015.
